# Performance evaluation of artificial neural network and multiple linear regression in the prediction of body mass index in children

**DOI:** 10.1101/2023.06.01.23290846

**Authors:** Muhammad Asif, Ghazi Khan Khosa, Abdullah Mohammad Alomair, Mohammad Ahmed Alomair, Muhammad Aslam, Muhammad Arslan, Muhammad Sanaullah, Justyna Wyszyńska

## Abstract

The body mass index (BMI) provides essential medical information related to body weight for the treatment and prognosis prediction of different diseases. The main goal of the present study was to evaluate the performance of artificial neural network (ANN) and multiple linear regression (MLR) model in the prediction of BMI in children. The data from a total of 5,964 children aged 5 to 12 years were included in study. Age, gender, neck circumference (NC), waist circumference (WC), hip circumference (HpC), and mid upper arm circumference (MUAC) measurements were used to estimate the BMI of children. The ANN and MLR were utilized to predict the BMI. The predictive performance of these methods was also evaluated. Gender-wise average comparison showed that median values of all the anthropometric measurements (except BMI) were significantly higher in boys as compared to girls. For the overall sample, the BMI prediction model was, 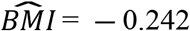 ― 0.147 X Age ― 0.367 X Gender + 0.176 X NC + 0.041 X WC + 0.060 X HpC + 0.404 X MUAC. A high R^2^ value and lower RMSE, MAPE, and MAD indicated that the ANN is the best method for predicting BMI in children. Our results confirm that the BMI of children can be predicted by using ANN and MLR regression methods. However, the ANN method has a higher predictive performance than MLR.

## Introduction

Evaluation of nutritional status is necessary for understanding the health of an individual or population. Children are considered a crucial population in regard to detecting their health status. This is especially important during infancy and adolescence, when the amount of water and adipose tissue undergo considerable changes[1-3]. Body composition changes are directly reflected in anthropometric measures, and the body mass index (BMI) is the most renowned and well-known criterion for determining nutritional status in both children and adults [4]. BMI is calculated by dividing weight in kilograms by height in square meters (i.e., kg/m^2^), and its cut-offs classify people as underweight, normal weight, overweight or obese [4]. Elevated BMI is strongly linked to the risk of developing cardiovascular health diseases (CHD), including diabetes, hypertension, dyslipidemia, and certain forms of cancer [5].

However, the measurements of height or weight could be difficult or even impossible due to the non-availability of accurate portable height scales, weighing machines, and well-trained health workers for collection of measurements. Accordingly, in similar scenarios, the BMI could not be measured. Therefore, the estimation of the BMI preferably to be a simple, affordable method, to be calculated with minimal equipment.

To date, various indirect methods have been developed to estimate BMI by measuring different body segments. Several studies [6-9] found that BMI correlates with neck circumference (NC), waist circumference (WC), hip circumference (HpC), and mid-upper arm circumference (MUAC), and Marshall et al. [10] found that measurement of WC, NC and MUAC can be used to estimate BMI accurately. Another study with Pakistani type 2 diabetes patients presented the prediction model for BMI based on WC and HpC measurements using multiple linear regression (MLR) analysis for the BMI prediction [11].

Over the last few years, several machine learning (ML) methods have also been applied to predict BMI or obesity efficiently. For example, a more recent study by Lee et al. [12] applied linear regression and different data mining algorithms (i.e., random forest and artificial neural network (ANN)) for the prediction of a newborn’s BMI based on ultrasound measures and maternal delivery information. Another study used k-nearest neighbor (KNN), classification and regression tree (CART), support vector machine (SVM), and multi-layer perceptron (MLP) algorithms, etc., to predict the BMI based on psychological variables [13]. The BMI of 1568 subjects aged 20-60 years was also predicted using three different data mining algorithms from voice signals information [14]. Sancar and Tabrizi [15] used an adaptive neuro fuzzy inference system (ANFIS) to calculate BMI based on metabolic risk factors. The existing literature has shown that no study considered age, gender, and different anthropometric measures as input variables in order to predict BMI in children, and performance of two different prediction methods i.e., ANN and MLR were rarely evaluated. This considerable research gap led to the conduction of this research study. The main goal of the study was to evaluate the performance of artificial neural network (ANN) and multiple linear regression (MLR) model in the prediction of BMI in children. The performance of ANN with MLR were evaluated by estimating the values of co-efficient of determination (R^2^), root mean square error (RMSE), mean absolute deviation (MAD) and mean absolute percentage error (MAPE).

## Materials and methods

A cross-sectional dataset collected from March to June 2016 multi-ethnic anthropometric survey (MEAS) was used in the present study. The data is also publicly available on the Mendeley website at https://data.mendeley.com/datasets/sxgymx5xjm/1. A detailed description of the study design, sampling methodology, and inclusion/exclusion criteria of the subjects in this survey has been described elsewhere [6, 16-18]. Briefly, in the MEAS, a total of 10,782 children and adolescents (aged 2-19 years) were recruited and the dataset of school-going children and adolescents (n= 9,929) aged 5 to 19 years was collected from 68 public and private schools. While data for subjects under the age of five were collected in public places such as markets, shopping malls and parks etc. This study only included 5,964 children aged 5 to 12 years. The raw dataset of different anthropometric measurements, i.e., body weight, height, NC, WC, HpC and MUAC, were taken in a comfortable standing position under standard procedure. The complete measurement protocols have been discussed in the previously published studies [6, 16-18]. From the height and weight measurements, BMI was calculated [BMI= weight (kg.) ÷ height (meters)^2^]. The dependent variable included in the study was the BMI of children while, age, gender of children and measurement of NC, WC, HpC and MUAC, were taken as independent variables.

The authors assert that the complete study was conducted in accordance with the ethical standards. The project was approved by the Institutional Ethics Research Board of Bahauddin Zakariya University, Multan under the registration number IRB# Stat-271/2017). Verbal informed consent was obtained from all participants and their parents. Verbal consent was witnessed and formally recorded. Researchers recorded on a form created specifically for documentation of verbal consent the name of the participant/parent who gave the verbal consent. The authors had access to information that could identify individual participants during or after data collection

### Statistical analysis

The entire statistical analysis was performed using SPSS ver. 23 (SPSS for Windows, Chicago, IL, USA). The normality of continuous variables (age, BMI, NC, WC, HpC and MUAC) was tested by using the Kolmogorov-Smirnoff test. The significance level of p < 0.05 was considered and the results were expressed as median [interquartile range (IQR) = Q_1_-Q_3_]. The Mann-Whitney U test was used for average comparisons between the groups. The Spearman’s rank correlation (r_s_) was used to investigate the correlation between BMI and other anthropometric measurements. Since a correlation of less than 0.3 (i.e., r < 0.30) is generally described as a weak correlation[19], therefore, the independent variables having a correlation of less than 0.3 with BMI, were excluded from the analysis. The significance level was set at α = 5% for the whole analysis.

For BMI prediction in children, two different methods, the ANN and MLR were applied. An ANN is a computing system consisting of simple inter-connected processing elements called neurons. The input signals (input data) pass through the network of neurons to generate the network response(s). Each neuron (except the input ones) receives information from several neurons through a connection in proportion to their weights, sums them up and modifies the sum through a non-linear transfer function before passing the signal to other neurons [20]. We also present the block diagram of the current study (Fig 1).

**Fig 1.** The study block diagram.

When modeling the neural network, a multi-layer perceptron with an input layer, hidden layer and an output layer was used. Initially, data on 5,964 children were divided into two different parts, i.e., training data and testing data. The training dataset consists of 70% of all the data (i.e., 4181) and the rest of the data (n=1783) is used as a testing phase. The input data were normalized before training the model. The network was trained in 5000 epochs for different numbers of neurons in the hidden layer. In each epoch, a training data set was selected randomly to prevent learning the especial order of data. The commonly used back-propagation training algorithm “scaled conjugate gradient (SCG)” was used for the training of the models. An activation transfer function called “hyperbolic-tangent” was used in all cases.

It is important to adjust the learning rate and momentum terms during the learning process of the neural networks. High weights may destroy the learning behaviour of neural networks. The learning rate is set at a small value to prevent the selection of high weights. Small learning rates slowdowns the learning process. Following Heydari et al. [21], the learning rate and momentum were set at 0.1 and 0.7. The input layer consists of 6 neurons corresponding to independent measures (age, gender, NC, WC, HpC and MUAC). Such measures were used to predict BMI in children. In the hidden layer, different numbers of neurons were used for the optimal selection of network architecture and to prevent overtraining.

The MLR analysis was also performed to predict BMI. A linear regression model that involves more than one predictor (regressor) variable is called an MLR model. In an MLR, the relationship between dependent (regressand) and more than one independent variable (s) is expressed by a linear regression equation. An MLR equation with *k* regressors, is given as under:

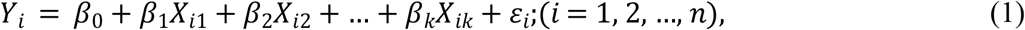

where

*Y*_*i*_ *= i*th observation of the dependent variable,

*X*_*ij*_ *= i*th observation of the *j*th regressor (*j* = 1, 2, …, *k*),

*β*_0_ *= Y*-intercept (the constant term),

*β*_*j*_ = regression coefficient corresponding to the *j*th regressor,

*ε*_*i*_ = the error term, assumed to be normal with zero mean and constant variance.

Referring to the MLR equation (1), in our study, the MLR equation would be

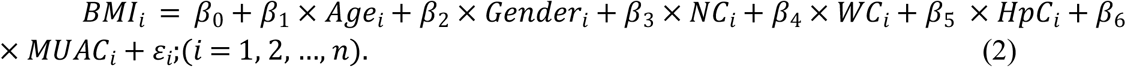

The gender of a child is coded as 1 for a boy and 0 for a girl.

The multicollinearity among the regressors was also determined using the variance inflation factor (VIF). The VIF between variables was < 5, suggesting that multicollinearity was not a problem in the models. For the evaluation of models’ prediction performance, different criteria, i.e., RMSE, MAPE, MAD and R^2^ were used in the literature [12, 13, 15]. For this study, the model prediction performance was also based on all the latter stated criteria. The model with the highest R^2^, lowest RMSE, MAPE and MAD was chosen as the final predictive model.

## Results

The study included 5,964 children (boys = 2865; 48.0% and girls = 3099; 52.0%) with a median age of 9.0 (IQR: 7.0-11.0). The descriptive statistics of anthropometric characteristics and Spearman’s correlation coefficients between BMI and NC, WC, HpC, and MUAC were listed (Table 1).

**Table 1.** Descriptive statistics and Spearman’s correlation coefficients between BMI and different anthropometric characteristics of the study subjects.

| Characteristics | Total (n=5964) | Boys (n=2865) | Girls (n=3099) | Correlation<br>($r_s$) |
| --- | --- | --- | --- | --- |
|  | Median (IQR) | Median (IQR) | Median (IQR) |  |
| BMI (kg/m<sup>2</sup>) | 15.26 (14.05-16.91) | 15.23 (14.09-16.76) | 15.26 (13.97-17.05)<sup>NS</sup> | 1.00 |
| NC (cm) | 25.40 (24.13-26.67) | 25.45 (24.13-26.67) | 25.40 (23.37-26.67)<sup>S</sup> | 0.55<sup>S</sup> |
| WC (cm) | 55.88 (50.80-60.96) | 56.39 (52.07-62.23) | 55.88 (50.80-60.96)<sup>S</sup> | 0.51<sup>S</sup> |
| HpC (cm) | 60.96 (55.88-67.31) | 63.50 (58.42-68.58) | 60.96 (55.88-66.04)<sup>S</sup> | 0.56<sup>S</sup> |
| MUAC (cm) | 16.51 (15.24-17.78) | 16.54 (15.24-17.79) | 16.51 (15.24-17.78)<sup>S</sup> | 0.63<sup>S</sup> |

IQR: Interquartile range; BMI: Body mass index; NC: Neck circumference; WC: Waist circumference; HpC: Hip circumference; MUAC: Mid-upper arm circumference. S: Significant < 0.001; NS: Not significant

A sex-based average comparison revealed that the median values of NC, WC, HpC and MUAC were significantly higher in boys than in girls. While the median BMI was not significantly different among the children of both sexes. In the study sample, significant positive correlations were observed between BMI and MUAC (r =0.63), followed by HpC (r =0.56), NC (r =0.56) and WC (r =0.51).

In order to predict the BMI values, the proposed MLR model (3) based on explanatory variables i.e., NC, WC, HpC, MUAC, age and gender (boys =1, girls=0) was used (Table 2).

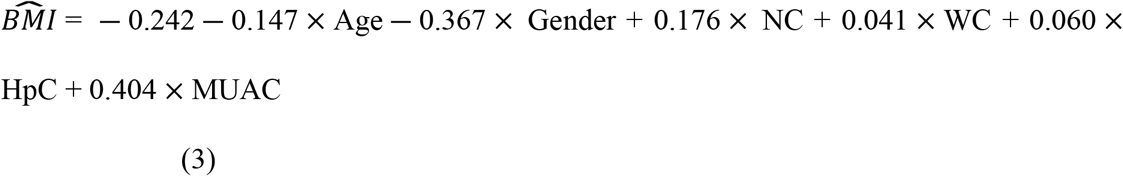

**Table 2.**
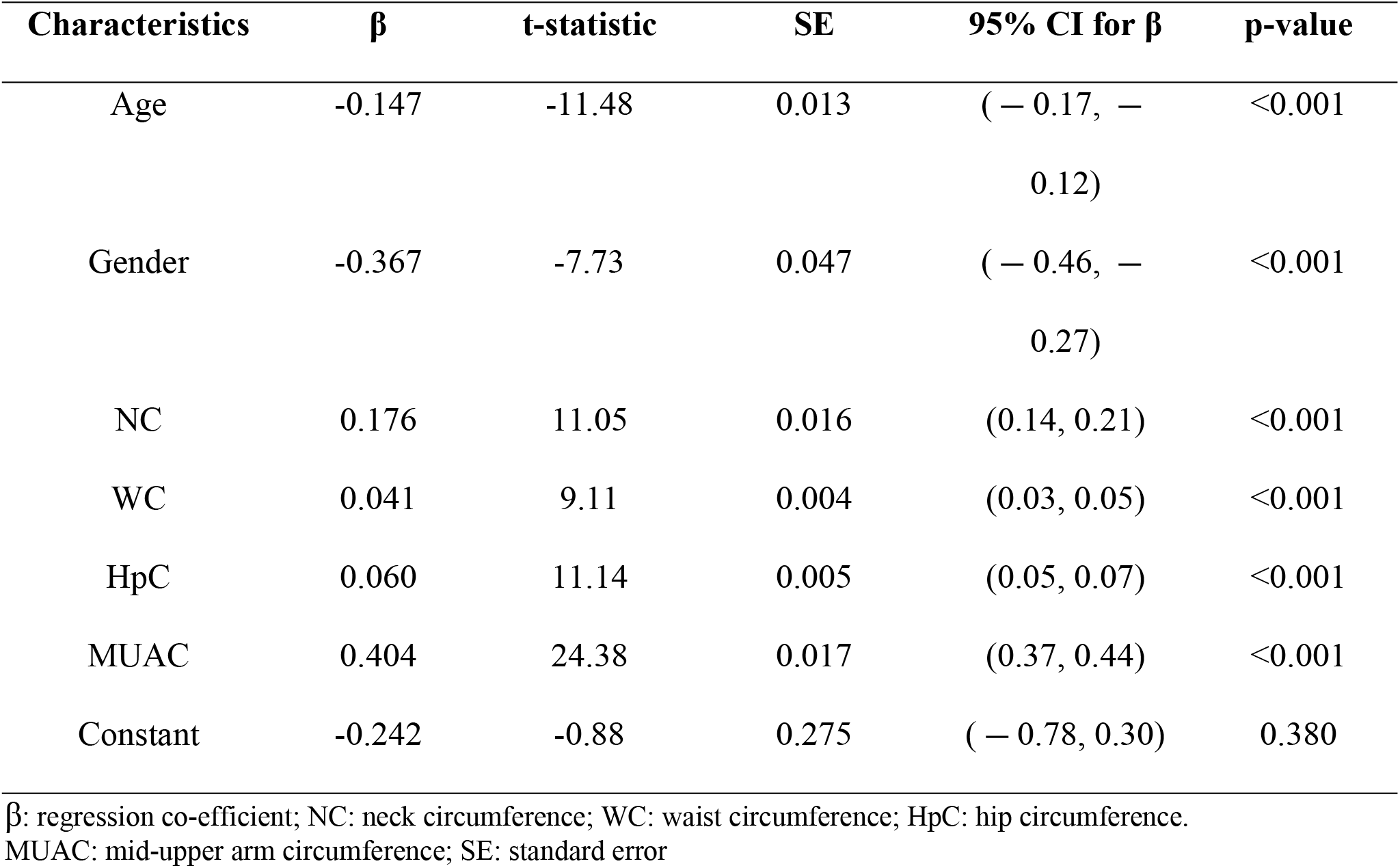
Estimated regression co-coefficients by using multiple linear regression.

For instance, the MLR model predicts a BMI value of 13.08 of a boy having age (5), NC (21.59), WC (50.80), HpC (48.26) and MUAC (13.97). Similarly, a BMI value of 14.97 was predicted for a girl having age (12), NC (26.67), WC (50.80), HpC (57.15) and MUAC (16.76). Moreover, an ANN model predicts the BMI values for the same boys and girls to be 13.13 and 15.52, respectively (see Table 3).

**Table 3.**
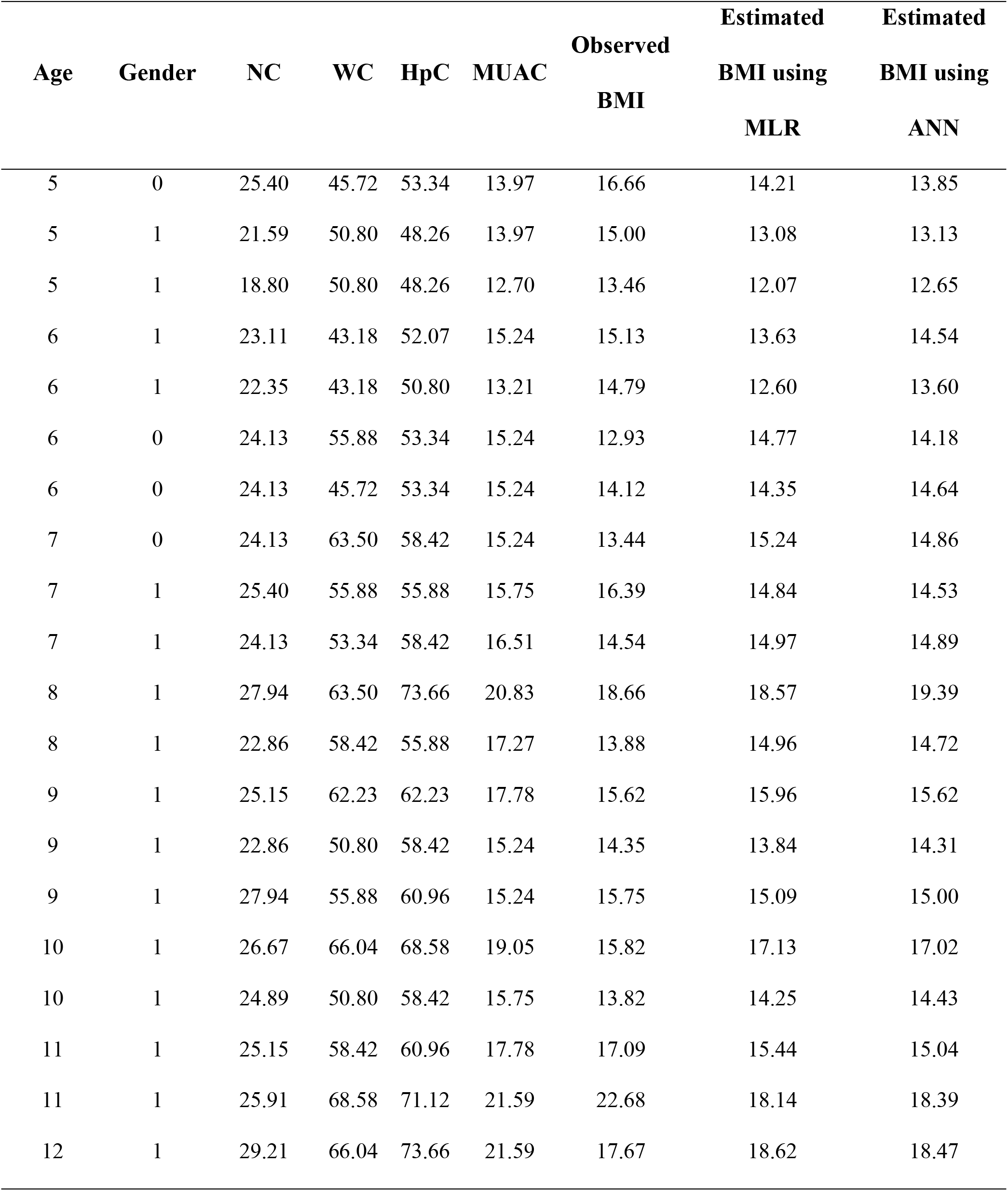
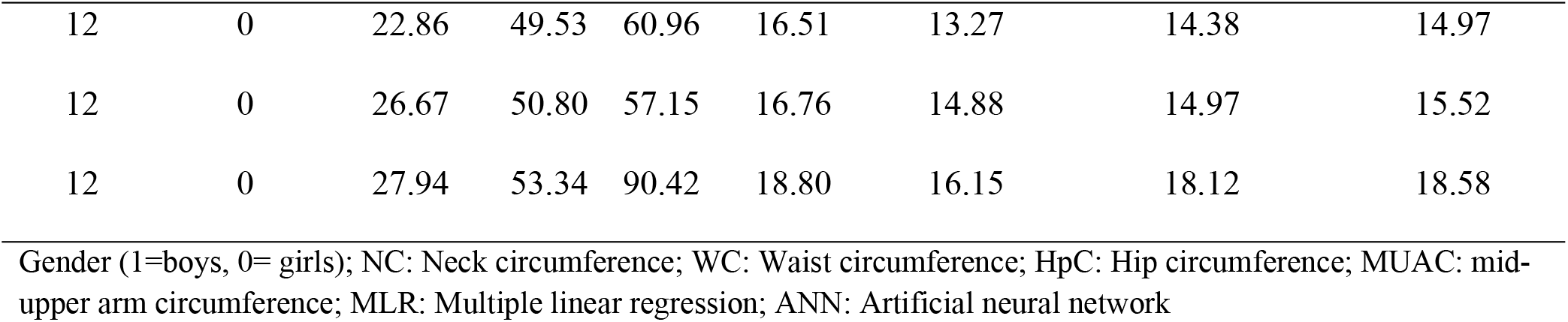
Comparison of observed and estimated BMI by using multiple linear regression and artificial neural network

| Age | Gender | NC | WC | HpC | MUAC | Observed<br>BMI | Estimated<br>BMI using<br>MLR | Estimated<br>BMI using<br>ANN |
| --- | --- | --- | --- | --- | --- | --- | --- | --- |
| 5 | 0 | 25.40 | 45.72 | 53.34 | 13.97 | 16.66 | 14.21 | 13.85 |
| 5 | 1 | 21.59 | 50.80 | 48.26 | 13.97 | 15.00 | 13.08 | 13.13 |
| 5 | 1 | 18.80 | 50.80 | 48.26 | 12.70 | 13.46 | 12.07 | 12.65 |
| 6 | 1 | 23.11 | 43.18 | 52.07 | 15.24 | 15.13 | 13.63 | 14.54 |
| 6 | 1 | 22.35 | 43.18 | 50.80 | 13.21 | 14.79 | 12.60 | 13.60 |
| 6 | 0 | 24.13 | 55.88 | 53.34 | 15.24 | 12.93 | 14.77 | 14.18 |
| 6 | 0 | 24.13 | 45.72 | 53.34 | 15.24 | 14.12 | 14.35 | 14.64 |
| 7 | 0 | 24.13 | 63.50 | 58.42 | 15.24 | 13.44 | 15.24 | 14.86 |
| 7 | 1 | 25.40 | 55.88 | 55.88 | 15.75 | 16.39 | 14.84 | 14.53 |
| 7 | 1 | 24.13 | 53.34 | 58.42 | 16.51 | 14.54 | 14.97 | 14.89 |
| 8 | 1 | 27.94 | 63.50 | 73.66 | 20.83 | 18.66 | 18.57 | 19.39 |
| 8 | 1 | 22.86 | 58.42 | 55.88 | 17.27 | 13.88 | 14.96 | 14.72 |
| 9 | 1 | 25.15 | 62.23 | 62.23 | 17.78 | 15.62 | 15.96 | 15.62 |
| 9 | 1 | 22.86 | 50.80 | 58.42 | 15.24 | 14.35 | 13.84 | 14.31 |
| 9 | 1 | 27.94 | 55.88 | 60.96 | 15.24 | 15.75 | 15.09 | 15.00 |
| 10 | 1 | 26.67 | 66.04 | 68.58 | 19.05 | 15.82 | 17.13 | 17.02 |
| 10 | 1 | 24.89 | 50.80 | 58.42 | 15.75 | 13.82 | 14.25 | 14.43 |
| 11 | 1 | 25.15 | 58.42 | 60.96 | 17.78 | 17.09 | 15.44 | 15.04 |
| 11 | 1 | 25.91 | 68.58 | 71.12 | 21.59 | 22.68 | 18.14 | 18.39 |
| 12 | 1 | 29.21 | 66.04 | 73.66 | 21.59 | 17.67 | 18.62 | 18.47 |
| 12 | 0 | 22.86 | 49.53 | 60.96 | 16.51 | 13.27 | 14.38 | 14.97 |
| 12 | 0 | 26.67 | 50.80 | 57.15 | 16.76 | 14.88 | 14.97 | 15.52 |
| 12 | 0 | 27.94 | 53.34 | 90.42 | 18.80 | 16.15 | 18.12 | 18.58 |

Gender (1=boys, 0= girls); NC: Neck circumference; WC: Waist circumference; HpC: Hip circumference; MUAC: mid-upper arm circumference; MLR: Multiple linear regression; ANN: Artificial neural network

The value of co-efficient of determination (R^2^) using MLR analysis revealed that about 48.0% variation in BMI is explained due to the predictor variables including the age and sex. While R^2^ using the ANN algorithm exhibited that more (53.4%) variation in BMI is explained due to the predictor variables. The RMSE, MAPE, and MAD values were also found to be lower in the ANN as compared to the MLR model. Bases on these findings, we can conclude that ANN outperforms MLR in predicting BMI in children (Table 4).

**Table 4.** Predictive performance of multiple linear regression and artificial neural network.

| **Algorithms** | **Model Evaluation Criteria** |  |  |  |
| --- | --- | --- | --- | --- |
|  | **R<sup>2</sup>** | **RMSE** | **MAPE** | **MAD** |
| ANN | 0.534 | 1.7047 | 0.0821 | 1.2758 |
| MLR | 0.4782 | 1.7953 | 0.0810 | 1.2761 |

ANN: Artificial neural network; MLR: Multiple linear regression; R<sup>2</sup>: Co-efficient of determination; RMSE: Root mean square error; MAPE: Mean absolute percentage error; MAD: Mean absolute deviation.

## Discussion

Obesity in children has now evolved into a severe public health issue, and its incidence has grown rapidly in recent years across the world [4]. Researchers used BMI as internationally accepted measure for defining overweight and obesity in both children and adults [22, 23]. Different studies in recent years have also utilized some other anthropometric measurements, i.e., WC, MUAC and NC for obesity screening purposes in children [7-9]. Because these measurements had a good correlation with BMI (r = ∼0.60 to ∼0.85). Based on receiver operating characteristics (ROC) curves analysis, the diagnostic ability of WC, MUAC and NC to detect children with overweight and obesity was very high i.e., areas under the curve (AUC) values between 75.0 % to 97.0 % [7-9]. Therefore, these measurements can be used to predict BMI in children. To the author’s knowledge, this is the first study that explores whether BMI in children can be predicted from anthropometric measurements by using MLR and ANN algorithms.

In this study, the neural network was designed using the information of 5,964 children living in different cities of Pakistan, including six input variables (age, gender, NC, WC, HpC and MUAC) and BMI as the output variable. A high R^2^ and lower RMSE, MAPE values indicated that ANN is the best method for predicting BMI in children than MLR. These findings are consistent with the earlier reports on the topic [24, 25]. In a study of 321 adult individuals in Iran, an RMSE (0.94) and R^2^ (0.890) using ANN were much better than MLR (RMSE= 1.31 and R^2^= 0.882) [24]. Another Iranian study estimated the BMI for 470 adult individuals and reported low RMSE values as compared to our results [25]. A high disparity in R^2^ and RMSE results may be due to the fact that they predicted the BMI for adult individuals based on different metabolic syndrome components (i.e., WC, SBP, DBP, FG, HDL and TG) and on different environmental and physical activity-related factors. However, our study predicted the BMI for children aged 5-12 years based on age, gender and anthropometric-related information as input variables. Some studies also employed the ANN method for obesity prediction e.g., an Iranian study with 414 adults found that ANN with an accuracy of 81.2% is a more efficient method for obesity prediction than logistic regression (accuracy = 80.2%) [21]. Another study compared the performance of ANN and logistic regression methods for obesity prediction among 82 individuals, and found that ANN performed better than logistic regression [26]. Using MLR analysis, we also found that NC, WC, HpC and MUAC are significant predictors for predicting BMI. These results are consistent with an earlier study by Marshall et al. [10], which reported that measurements of WC, NC and AC can be used to accurately estimate BMI and another study with 24,485 Pakistani type 2 diabetes patients aged 20 years and above also offered a BMI prediction model based on WC and HpC measurements [11].

The major strength of the study is that we predicted the BMI in children based on NC, WC, HpC and MUAC whose measurements are simple, quick, and just require a non-stretchable plastic tape. Our recommendation is to include the metabolic risk-related variables like SBP, DBP, FG, LDL, HDL and TG that affect BMI. Some studies predicted the BMI by using voice signals information [14], psychological [13], environmental and physical activity-related variables [25], and it would be an important contribution to extend this study based on these new variables. Lastly, the focus of this study was to predict BMI for children and however BMI estimation for adult individuals need to be studied and it will be a good topic for future research. However, the study has limitations. Since our study has a cross-sectional character, we cannot conclude a cause-and-effect relationship. The limitation of this study was a single anthropometric measurements, thus intra-observer variability could not be calculated. However, these measurement was always performed by the same researcher and there was no inter-observer variability between measurements.

## Conclusion

The findings of this study imply that both methods, MLR and ANN can be used to predict BMI in children. The use of ANN, a high R^2^ and lower RMSE, MAPE and MAD values demonstrate that this method is biologically acceptable and more effective for predicting BMI based on age, gender and four different anthropometric variables. Our methods and results can be used for obesity prediction in Pakistani children as an alternative to the clinical findings and public health research. Further research to overcome the present study’s limitations is also required.

## Data Availability

The dataset can be accessed through the following link: https://data.mendeley.com/datasets/sxgymx5xjm/1

## Acknowledgments

The author(s) are thankful to Mr. Muhammad Qasim, who reviewed the statistical interpretation and made corrections if required.

**The author (s) received no specific funding for this work**.

## Notes

### Competing Interest Statement

The authors have declared no competing interest.

### Author Declarations

The project was approved by the Institutional Ethics Research Board of Bahauddin Zakariya University, Multan under the registration number IRB# Stat-271/2017). Verbal informed consent was obtained from all participants and their parents. Verbal consent was witnessed and formally recorded.

## References

1. Mihalopoulos NL, Holubkov R, Young P, Dai S, Labarthe DR. Expected changes in clinical measures of adiposity during puberty. J Adolesc Health 2010; 47:360–66.

2. Talwar I, Sharma K, Kapur S. Growth trends in body, fat, circumferential and physiological traits during adolescents among Rajput females of Theog, Shimla District, India. Ann Hum Biol 2010; 37:536–53.

3. Wells JC, Fuller NJ, Dewit O, Fewtrell MS, Elia M, Cole TJ. Four component model of body composition in children: density and hydration of fat-free mass and comparison with simpler models. Am J Clin Nutr 1999; 69:904–12.

4. WHO Consultation on Obesity (1999: Geneva, Switzerland) & World Health Organization. (2000). Obesity : preventing and managing the global epidemic : report of a WHO consultation. World Health Organization. https://apps.who.int/iris/handle/10665/42330

5. Field AE, Coakley EH, Must A, Spadano JL, Laird N, Dietz WH, et al. Impact of overweight on the risk of developing common chronic diseases during a 10-year period. Arch Int Med 2001; 161(13):1581–86.

6. Asif M, Aslam M, Mustafa S, Alfat S. Age-specific Differences and Interrelations between Anthropometric Variables in Pakistani Children aged 2 to 19 years. Rawal Med J 2018; 43:164–69.

7. Nafiu OO, Burke C, Lee J, Voepel-Lewis T, Malviya S, Tremper KK. Neck circumference as a screening measure for identifying children with high body mass index. Pediatrics 2010; 126(2):e306–10.

8. Mazicioglu MM, Hatipoglu N, Oztürk A, Ciçek B, Ustünbas HB, Kurtoglu S. Waist circumference and mid-upper arm circumference in evaluation of obesity in children aged between 6 and 17 years. J Clin Res Pediatr Endocrinol 2010; 2:144–50.

9. Asif M, Aslam M, Altaf, S. Mid-upper-arm circumference as a screening measure for identifying children with elevated body mass index: a study for Pakistan. Korea J Pediatr 2018; 61:6–11.

10. Marshall AR, Haboubi N, Jones S. Body mass index estimation from waist, neck and mid-arm circumference. Gastrointestinal Nurs 2011; 9:37–40.

11. Ghias M, Khawaja KI, Masud F, Atiq S, Khalid Pervaiz M. A new approach for estimation of body mass index using waist and hip circumference in type 2 diabetes patients. J Ayub Med Coll Abbottabad 2010; 22:111–116.

12. Lee KS, Kim HY, Lee SJ, Kwon SO, Na S, Hwang HS, et al. Korean Society of Ultrasound in Obstetrics and Gynecology Research Group. Prediction of newborn’s body mass index using nationwide multicenter ultrasound data: a machine-learning study. BMC Pregnancy Childbirth 2021; 21(1):172.

13. Delnevo G, Mancini G, Roccetti M, Salomoni P, Trombini E, Andrei F. The Prediction of Body Mass Index from Negative Affectivity through Machine Learning: A Confirmatory Study. Sensors (Basel) 2021; 21(7):2361.

14. Lee BJ, Kim KH, Ku B, Jang JS, Kim JY. Prediction of body mass index status from voice signals based on machine learning for automated medical applications. Artif Intell Med 2013; 58(1):51–61.

15. Sancar N, Tabrizi SS. Body mass index estimation by using adaptive neuro fuzzy inference system. Procedia Comput Sci 2017; 108C:2501–06.

16. Asif M, Aslam M, Qasim M, Altaf S, Ismail A, Ali H. A dataset about anthropometric measurements of the Pakistani children and adolescents using a cross-sectional multi-ethnic anthropometric survey. Data Brief 2020; 34:106642.

17. Asif M, Aslam M, Altaf S, Mustafa S. Developing waist circumference, waist-to-height ratio percentile curves for Pakistani children and adolescents aged 2-18 years using Lambda-Mu-Sigma (LMS) method. J Pediatr Endocrinol Metab 2020; 33:983–93.

18. Asif M, Aslam M, Khan S, Altaf S, Ahmad S, Qasim M, Ali H, Wyszynska J. Developing neck circumference growth reference charts for Pakistani children and adolescents using the lambda-mu-sigma and quantile regression method. Public Health Nutr 2021; 24(17):5641–49.

19. Profilldis VA, and Botzoris GN. Modeliing of Transport Demand, Analyzing, Calculating, and Forecasting Transport Demand. Ch # 5, 2019, pp. 163-224. DOI: https://doi.org/10.1016/B978-0-12-811513-8.00005-4.

20. Dreyfus G. Neural networks: methodology and applications. Springer Verlag, 2005. DOI: 10.1007/3-540-28847-3.

21. Heydari ST, Ayatollahi SM T, Zare N. Comparison of Artificial Neural Networks with Logistic Regression for Detection of Obesity. J Med Syst 2012; 36:2449–54.

22. Ogden CL, Carroll MD, Flegal KM. High body mass index for age among US children and adolescents, 2003-2006. JAMA 2008; 299:2401–5.

23. Lobstein T, Baur L, Uauy R. Obesity in children and young people: a crisis in public health. Obes Rev 2004, 5,4–85.

24. Tabrizi SS, Sancar N. Prediction of Body Mass Index: A comparative study of multiple linear regression, ANN and ANFIS models. Procedia Com Sci 2017; 120:394–401.

25. Hoseini SH, Soltani A, Pourahmadi-Nakhli M. Application of artificial neural network (ANN) in estimation of body mass index (BMI) based on the connection between environmental factors and physical activity. Int J Artif Intell 2012; 3:107.

26. Ergün U. The classification of obesity disease in logistic regression and neural network methods. J Med Syst 2009; 33:67–72.

